# Mental Health Outcomes of Foster and Adopted Individuals with Adverse Childhood Experiences: A Validation of Known Risks Using EHR Data

**DOI:** 10.64898/2026.05.28.26354276

**Authors:** Anita C Randolph, Evan Dastin-van Rijn, Shelby Anderson, Logan Caola, Erich Kummerfeld, Christi Sullivan, Stefanie Simpson, Aarav Kallar, Ritwick Banerjee, Audrey Houghton, Mark Fiecas

**Author notes:** **Corresponding Author:** Audrey Houghton, BA **Corresponding Author Address:** 2312 S. 6th St. Floor 2, Suite F221 Minneapolis, MN 55454 **Corresponding Author Email:**.

## Abstract

**Background:** Adverse childhood experiences (ACEs) are traumatic or adverse events in early life that can have lasting effects on behavioral, emotional, and psychological functioning. Prior research suggests ACEs relate to later psychiatric outcomes through threshold, cumulative, and individual-specific risk patterns. Few studies, however, have operationalized all three models to test ACE-specific associations with diagnosed psychiatric disorders in individuals who are adopted or with foster care histories.

**Methods:** We conducted a cross-sectional retrospective study using electronic health record data from foster care and adopted patients aged 0-21 years old seen at the University of Minnesota Adoption Medicine Clinic (UMN-AMC) between 2014-2024. Extracted measures included ACE history, demographics, and psychiatric diagnoses. We used latent class analysis and logistic regression to identify clusters of adversity and estimate associations with psychiatric diagnosis domains, adjusting for Sex and Age at Initial Visit.

**Results:** ACEs showed a threshold pattern across psychiatric domains, with higher ACE counts associated with greater odds of psychiatric diagnoses. Individual risk modeling indicated that exposure to abuse or violence was associated with higher odds of psychiatric diagnoses. Across cumulative and individual risk approaches, Anxiety Disorders, Mood Disorders, and Behavioral or Emotional Disorders showed the greatest sensitivity to adversity.

**Conclusion:** Current ACE models may not fully capture neurodevelopmental impacts reflected in diagnosed psychiatric disorders among adolescents, particularly in high-risk groups such as foster and adopted individuals. In a large clinic sample our findings support a nuanced association between ACEs and later psychiatric diagnoses and highlight the need for ACE-focused assessment, prevention, and treatment strategies tailored to foster care and adopted populations.

## Background

In the U.S., two-thirds of adults have experienced at least one adverse childhood experience (ACE), and one in six have experienced four or more ACEs. Understanding how ACEs impact future health outcomes is a growing area of research.^1^ ACEs are traumatic events occurring during the first 18 years of a person’s life, and include neglect, physical/sexual/emotional abuse, and household dysfunction like caregiver mental illness or substance use. Risk factors for ACE exposure include housing instability, community-level adversity (e.g., violence, food insecurity), and intergenerational trauma.^2^ The relationship between ACEs and poor adult mental health outcomes are well-documented across psychiatric disorders,^3^ including evidence that ACE exposure increases the odds of multiple mood and anxiety disorders.^4,5^ ACEs are also associated with health-harming behaviors in adulthood, underscoring their public health impact.^6^

Over the past three decades, several frameworks have been used to characterize ACE-related risk. Threshold (dose-dependent) models emphasize higher ACE counts as conferring greater risk of adverse outcomes,^3,7^ including markedly elevated prevalence of depression, substance use, and suicide attempts amongst those with ≥4 ACEs.^8^ Cumulative risk models sum exposures into a weighted score to identify individuals at highest risk,^9^ but often assume equal weighting and can obscure heterogeneity in co-occurring adversities and ACE-specific drivers of outcomes.^10^ In contrast, individual risk models estimate associations for specific ACE types, supporting more targeted prevention strategies.^11^ For example, sexual abuse, emotional abuse, and community violence have each been linked to increased risk for clinically confirmed psychiatric disorders.^12^

Clarifying how these risk models operate in high-risk groups such as foster care and adopted individuals is essential for prevention and early intervention. Higher ACE burden in the foster care system has been associated with poorer permanency outcomes, and foster care-involved youth have substantially elevated mental health needs relative to peers.^13^ We investigated threshold, cumulative, and individual ACE risk models in relation to psychiatric diagnoses among foster care and adopted patients receiving care at the University of Minnesota’s Adoption Medicine Clinic (UMN-AMC). We hypothesized that higher ACE exposure would be associated with increased odds of psychiatric diagnosis, with additional ACE-type specificity across diagnostic groupings.

## Methods

### Study Participants

The study population was patients with available ACE and psychiatric diagnoses data via patient UMN-AMC electronic health records (EHRs). The UMN-AMC serves children who experience higher rates of early-life adversities than the general population, including children currently in the foster care system, those recently adopted domestically and internationally, and adopted children with severe milestone delays and mental health disorders. Because our sample spans from newborn to 21 years of age, we describe our sample using the word "individual" in place of words like "children" or "youth" to best represent the age span of our population.

### Design and data extraction

Data was collected from the EHRs of the UMN-AMC of 3,685 patients ages 0-21 years old who completed their first Comprehensive Child Wellness Assessments (CCWA) from 2014 through 2024. Five trained Research Professionals extracted ACE data from narrative notes provided by social workers, child protective services, and relatives during the CCWA visit. Psychiatric diagnoses were pulled from each patient’s EHR by Best Practices Integrated Informatics Core (BPIC) staff. Patients were excluded from analyses if their EHR data were inaccessible or incomplete (eg, for patients who either did not attend or schedule their follow up appointment(s) or new to the clinic and had no historic clinical data from 2014-2024). This resulted in a final analytical sample of n = 2720.

## Measures

### Adverse Childhood Experiences Questionnaire

The 15-item ACE questionnaire uses expanded definitions based on the Kaiser-CDC 10-item ACE questionnaire covering abuse (physical, emotional, sexual), neglect (physical, emotional), and household dysfunction (substance use, mental illness, maternal violence, and criminal behavior).^9^ ACE definitions were further expanded using the PEARLS screener to capture bullying, neighborhood violence, discrimination, housing instability, and food insecurity. See Table 1 for collected variables and classification.

**Table 1.** Definitions and operationalization of ACEs questionnaire.

| ACE | Definition | Operationalization |
| --- | --- | --- |
| Emotional abuse | Has a parent/caregiver ever sworn, insulted, humiliated, or put down your child? | Yes/no/suspected |
| Physical abuse | Has any adult in the household ever pushed, grabbed, slapped, or thrown something at your child, hit your child so hard that your child had marks or was injured, threatened your child in a way that made your child afraid that they might be hurt? | Yes/no/suspected |
| Sexual abuse | Has your child ever experienced sexual abuse?<br>For example, anyone touched your child or asked your child | Yes/no/suspected |
|  | to touch that person in a way that was unwanted, or made your child feel uncomfortable, or anyone ever attempted or actually had oral, anal, or vaginal sex with your child, or allowed to witness adult sexual activity. |  |
| Abuse unspecified | This variable captures any abuse mentioned but not specified as emotional, physical, or sexual within the patient's EHR. | Yes/no/suspected |
| Physical neglect | Has your child ever lacked appropriate care by any caregiver? Examples include not being protected from unsafe situations, not cared for when sick or injured even when the resources were available, not having enough food to eat, forced to wear dirty clothes, caregiver being unable to take child to doctor if needed, and or abandoned. | Yes/no/suspected |
| Neglect unspecified | This variable captures any neglect mentioned in the EHR not specified as physical neglect. Examples include emotional, educational, and medical neglect. | Yes/no/suspected |
| Caregiver treated violently | Has your child ever seen or heard a parent/caregiver being slapped, kicked, punched, beat up, pushed, bitten, hit with a fist, hit with something hard, threatened, or hurt with a weapon, had something thrown at, sexually assaulted or raped, screamed at, sworn at, insulted or humiliated by another adult/household | Yes/no/suspected |
|  | member? |  |
| Family violence | Has your child ever seen or heard a non-caregiver being slapped, kicked, punched, beat up, pushed, bitten, hit with a fist, hit with something hard, threatened, hurt with a weapon, had something thrown at them, repeatedly hit over at least a few minutes, screamed at, sworn at, insulted or humiliated, sexually assaulted or raped by another adult/household member?<br>or<br>This includes another category classifying if the violence is perpetrated by the patient. Has your child ever slapped, kicked, punched, beat up, pushed, bitten, hit with a fist, hit with something hard, threatened, or hurt with a weapon a non-caregiver family member | Yes/no/suspected/perpetrated by patient |
| Substance abuse in the household | Has the child's caregiver ever had, or currently has a problem with too much alcohol, street drugs, or prescription medication use? | Yes/no/suspected |
| Mental Illness in the household | Has the child ever lived with a parent/caregiver who had mental health issues or attempted suicide? | Yes/no/suspected |
| Incarcerated Household Member | Has the child ever lived with a parent/caregiver who went to jail/prison? | Yes/no/suspected |
| Parental Divorce | Have there ever been significant changes in the relationship status of the | Yes/no/suspected |
|  | child's caregiver? |  |
| Caregiver Disability | Has the child ever lived with a parent/caregiver who had a serious physical illness or disability (physical, intellectual, learning, developmental, or sensory)? | Yes/no/suspected |
| Housing Instability | Has the child ever had problems with housing? (for example, being homeless, not having a stable place to live, faced eviction or foreclosure, staying in a shelter, having to move frequently) | Yes/no/suspected |
| Community Violence | Has your child ever perpetuated, seen, heard, or been a victim of violence in your neighborhood? | Yes/no/suspected/perpetrated by patient |
| Personal history of being arrested, detained, or incarcerated | Has your child ever been detained, arrested or incarcerated? | Yes/no/suspected |
| Personal history of suicidal ideation or attempt | Has your child ever had personal history of suicidal ideation or attempt? | Yes/no/suspected |
| Personal history of self injurious behavior | Has your child ever had a personal history of self injurious behavior? | Yes/no/suspected |
| Personal history of substance use | Has your child ever had a personal history of substance use? | Yes/no/suspected |

### Psychiatric disorders

We used ICD-10 diagnostic codes to identify psychiatric diagnoses recorded at visits. ICD-10 is the World Health Organization’s classification system for health conditions; codes include a letter and numbers (e.g., generalized anxiety disorder: F41.1), where the initial letter denotes the diagnostic chapter (F = mental, behavioral, and neurodevelopmental disorders). ICD-10 codes were extracted from patients’ EHRs via the UMN BPIC, which compiled each participant’s medical and psychiatric diagnoses across their record. Because some earlier visits used ICD-9, both ICD-9 and ICD-10 codes were included in analyses. Diagnostic outcome operationalization is provided in Table 2.

**Table 2.** Mapping of diagnostic categories to ranges of ICD codes.

| Diagnosis Category | ICD9 Codes | ICD10 Codes |
| --- | --- | --- |
| **Mood Disorders** | 311 | F30-F39 |
| **Intellectual and Developmental Disorders** | 315, 317-319 | F80-F84, F70-F79 |
| **Anxiety Disorders** | 300, 308-309 | F40-F48 |
| **Substance Use Disorders** | 304-305, 291-292 | F10-F19 |
| **Behavioral and Emotional Disorders** | 312-314 | F90-F98 |
| **Personality Disorders** | 301 | F60-F69 |
| **Behavioral Syndromes Associated with Physiological Disturbances** | 302, 306-307 | F50-F59 |

## Statistical Analysis

Diagnosis codes were grouped by the first three ICD characters and then collapsed into broader diagnostic categories (Table 2). Each category served as a binary outcome in logistic regression (1 = ever diagnosed; 0 = never diagnosed during the observation period). For each outcome, we fit three models to estimate marginal associations: (1) categorical ACE count (1, 2, 3, ≥4 vs 0), (2) cumulative ACE score (capped at 10 to limit influence of rare extremes; less than 2% of the individuals had more than 10 ACEs.), and (3) univariate associations for individual ACE types or personal history metrics. All models adjusted for Sex and Age at Initial Visit. P-values were false discovery rate-controlled at 0.05 using Benjamini–Hochberg. Regression coefficients and 95% confidence intervals (CIs) were converted to odds ratios (ORs); for cumulative score, ORs are reported per one SD increase to aid comparability with categorical predictors.

## Results

Results from descriptive measures are presented in Table 3. Among the mental health diagnoses assessed in the study, the most prevalent were Anxiety Disorders (60.3%) and Intellectual and Developmental Disorders (47.9%), and the least prevalent were Substance Use Disorders (4.9%) and Personality Disorders (3.9%). ACEs were highly prevalent in the dataset, with 80.1% of the individuals having at least one type of adverse experience, and 47.2% having four or more. Among ACEs, substance abuse in the household (49.9%) and mental illness in the household (42.8%) were the most prevalent, while family violence (11.8%) and unspecified types of abuse (5.1%) were the least prevalent. A sizable minority of the individuals also had personal experience with health-harming behaviors, among which personal history of self injurious behavior (18.9%) was the most prevalent while personal history of substance use (2.7%) was the least prevalent.

**Table 3.**
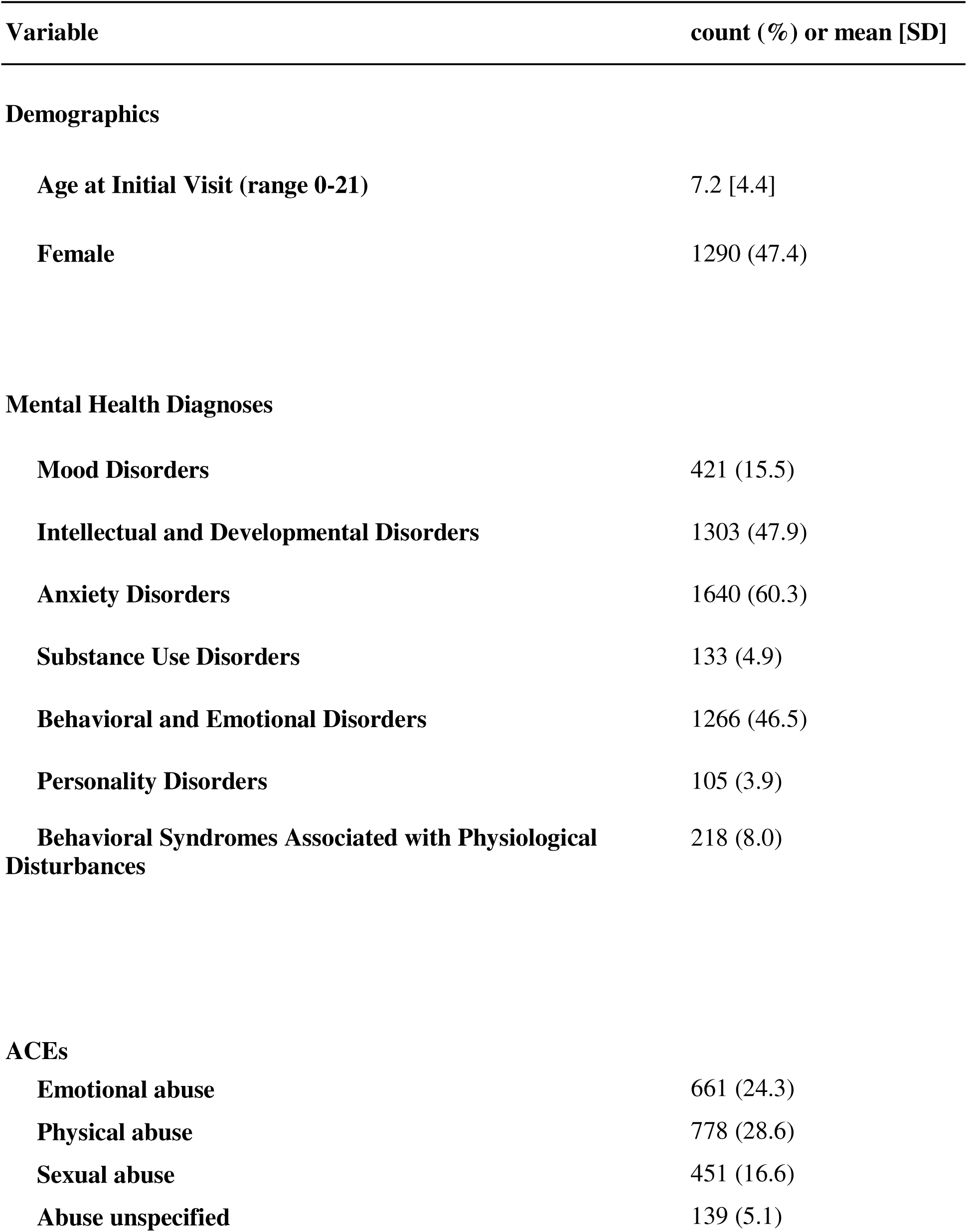
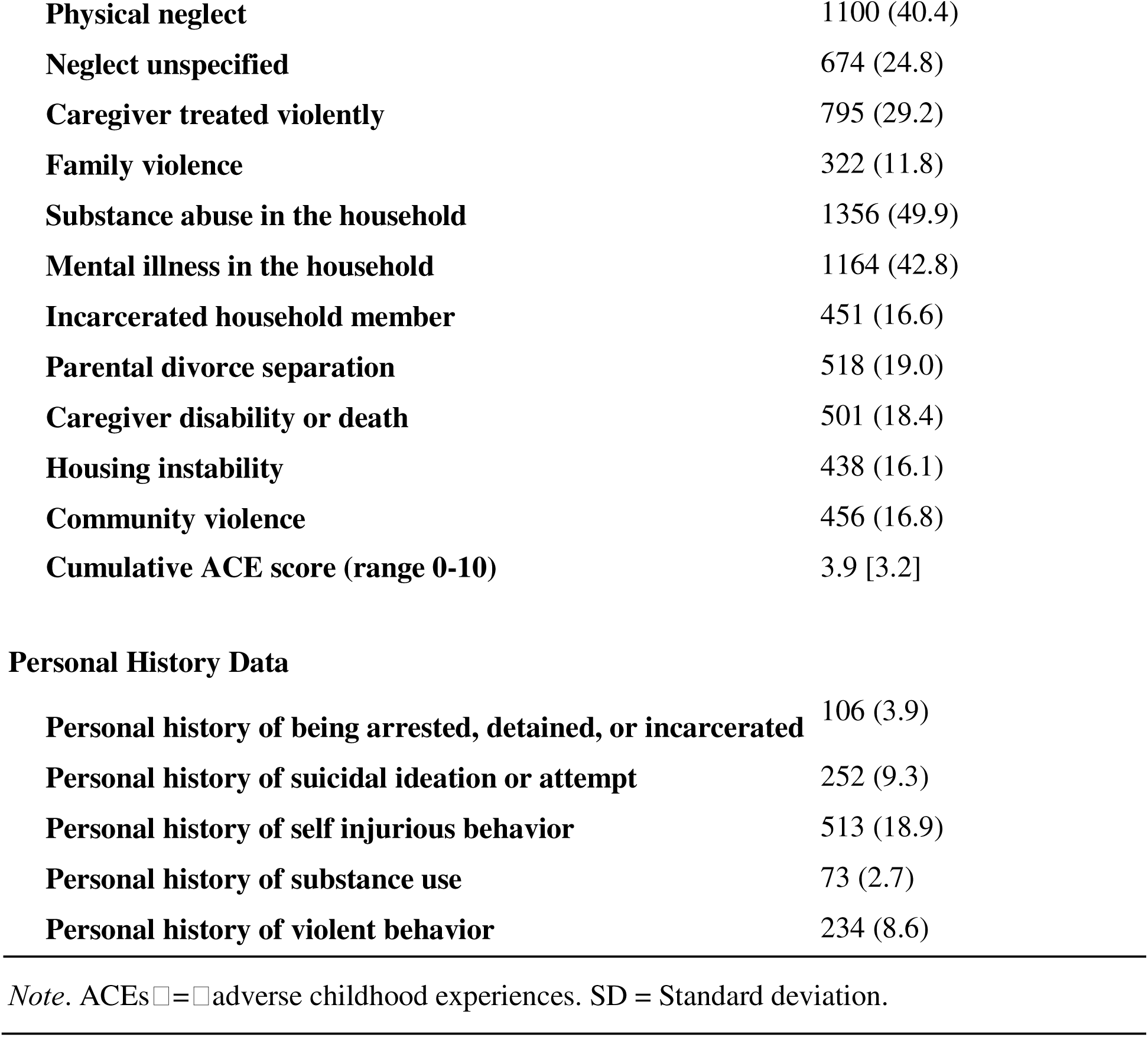
Description of study measures (N=2720)

| **Variable** | **count (%) or mean [SD]** |
| --- | --- |
| **Demographics** |  |
| **Age at Initial Visit (range 0-21)** | 7.2 [4.4] |
| **Female** | 1290 (47.4) |
| **Mental Health Diagnoses** |  |
| **Mood Disorders** | 421 (15.5) |
| **Intellectual and Developmental Disorders** | 1303 (47.9) |
| **Anxiety Disorders** | 1640 (60.3) |
| **Substance Use Disorders** | 133 (4.9) |
| **Behavioral and Emotional Disorders** | 1266 (46.5) |
| **Personality Disorders** | 105 (3.9) |
| **Behavioral Syndromes Associated with Physiological Disturbances** | 218 (8.0) |
| **ACEs** |  |
| **Emotional abuse** | 661 (24.3) |
| **Physical abuse** | 778 (28.6) |
| **Sexual abuse** | 451 (16.6) |
| **Abuse unspecified** | 139 (5.1) |
| **Physical neglect** | 1100 (40.4) |
| **Neglect unspecified** | 674 (24.8) |
| **Caregiver treated violently** | 795 (29.2) |
| **Family violence** | 322 (11.8) |
| **Substance abuse in the household** | 1356 (49.9) |
| **Mental illness in the household** | 1164 (42.8) |
| **Incarcerated household member** | 451 (16.6) |
| **Parental divorce separation** | 518 (19.0) |
| **Caregiver disability or death** | 501 (18.4) |
| **Housing instability** | 438 (16.1) |
| **Community violence** | 456 (16.8) |
| **Cumulative ACE score (range 0-10)** | 3.9 [3.2] |
| Personal History Data |  |
| **Personal history of being arrested, detained, or incarcerated** | 106 (3.9) |
| **Personal history of suicidal ideation or attempt** | 252 (9.3) |
| **Personal history of self injurious behavior** | 513 (18.9) |
| **Personal history of substance use** | 73 (2.7) |
| **Personal history of violent behavior** | 234 (8.6) |

*Note.* ACEs = adverse childhood experiences. SD = Standard deviation.

Table 4 presents the results from logistic regressions testing relations between categorical ACE count and neuropsychiatric outcomes. Modeling showed that increasing ACE counts were associated with increased odds of poor neuropsychiatric outcomes across diagnoses with the exception of Intellectual and Developmental Disorders. To illustrate, compared to individuals with no ACEs, the odds of receiving an Anxiety Disorder diagnosis was 0.99 (CI = 0.77-1.28) with one ACE, 1.60 (CI = 1.18-2.17) with two ACEs, 1.80 (CI = 1.30-2.48) with three ACEs, and 3.48 (CI = 2.79-4.34) with four or more ACEs. A second set of regression models also indicated that weighted ACE score was associated with Mood Disorders (OR = 1.26, CI = 1.13-1.40), Anxiety Disorders (OR = 1.93, CI = 1.76-2.12), Behavioral and Emotional Disorders (OR = 1.33, CI = 1.23-1.44), Personality Disorders (OR = 1.34, CI = 1.10-1.63), and Behavioral Syndromes Associated with Physiological Disturbances (OR = 1.18, CI = 1.02-1.35). Effect sizes and confidence intervals for all ACE count and score coefficients are visualized in Figure 1A.

**Figure 1:**
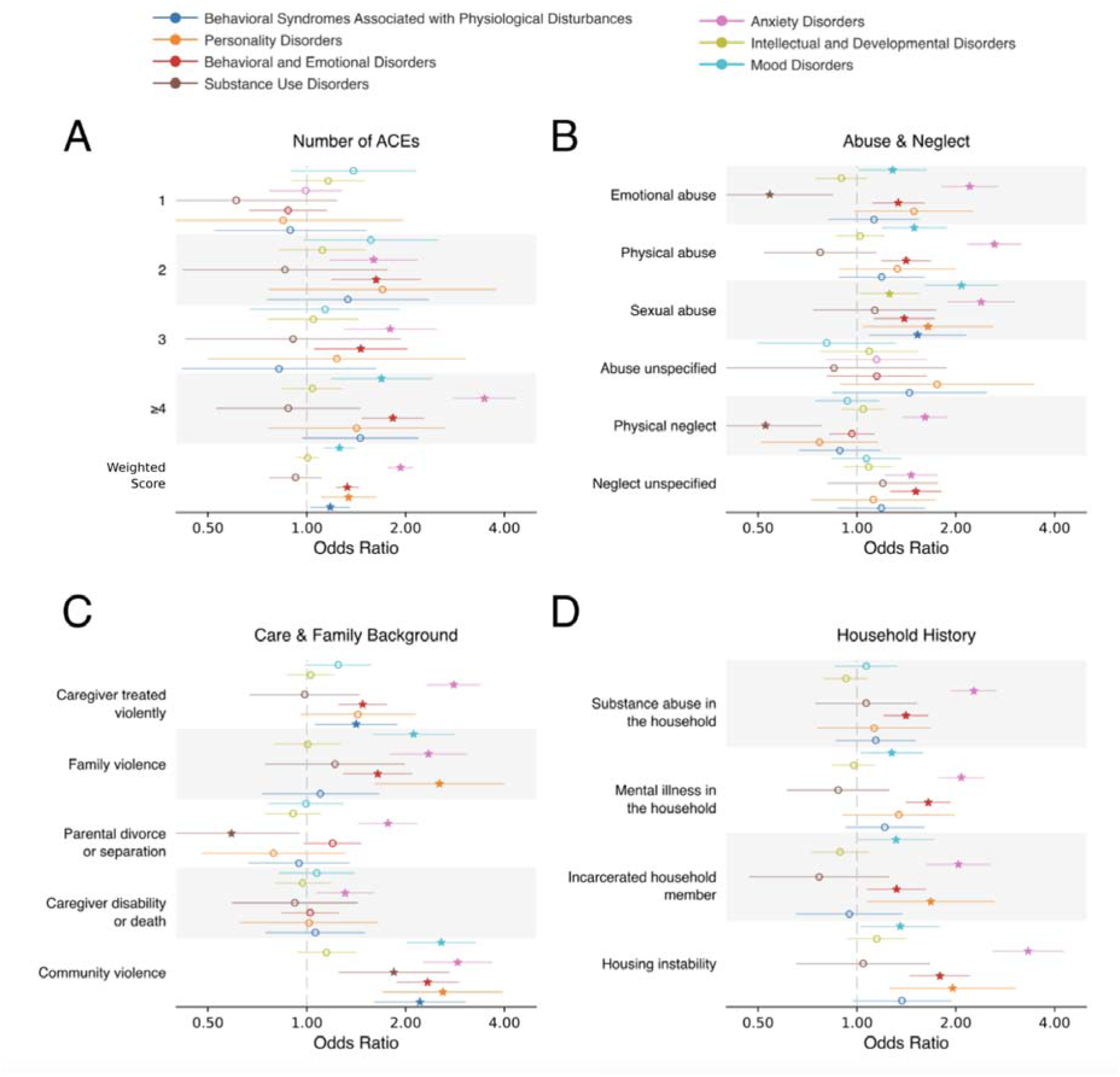
Forest plot of marginal effects of ACE counts and categories on psychiatric indications. Each panel shows estimated effects and 95% confidence intervals for ACE count and categories across seven diagnostic outcomes. Points indicate mean parameter estimates for odds ratios, and horizontal lines denote corresponding confidence intervals. Significant predictors are indicated by star markers. Outcomes for each ACE feature are grouped by row. Panels correspond to different groupings of ACE features (**A** - Number of ACEs, **B** - ACEs related to abuse & neglect, **C** - ACEs related to care and family background, and **D** - ACEs related to personal and household history). For the ACE score, the odds ratio corresponds to a one standard deviation increase.

**Table 4.** Associations between ACE counts and neuropsychiatric outcomes. Threshold Risk Model = 1, 2, 3, ≥4 exposures; Cumulative Score Risk Model = Weighted Score.

| **Diagnosis** | **Number of ACEs** | **OR (95 % CI)** |
| --- | --- | --- |
| **Mood Disorders** | 1 | 1.39 (0.90, 2.15) |
|  | 2 | 1.57 (0.98, 2.51) |
|  | 3 | 1.14 (0.67, 1.93) |
|  | ≥4 | 1.69 (1.19, 2.41)** |
|  | Weighted Score (0-10) | 1.26 (1.13, 1.40)*** |
| **Intellectual and Developmental Disorders** | 1 | 1.16 (0.90, 1.50) |
|  | 2 | 1.11 (0.82, 1.51) |
|  | 3 | 1.05 (0.76, 1.44) |
|  | ≥4 | 1.04 (0.84, 1.28) |
|  | Weighted Score (0-10) | 1.01 (0.93, 1.09) |
| **Anxiety Disorders** | 1 | 0.99 (0.77, 1.28) |
|  | 2 | 1.60 (1.18, 2.17)** |
|  | 3 | 1.80 (1.30, 2.48)*** |
|  | ≥ 4 | 3.48 (2.79, 4.34)*** |
|  | Weighted Score (0-10) | 1.93 (1.76, 2.12)*** |
| **Substance Use Disorders** | 1 | 0.61 (0.30, 1.24) |
|  | 2 | 0.86 (0.42, 1.76) |
|  | 3 | 0.91 (0.43, 1.93) |
|  | ≥ 4 | 0.88 (0.53, 1.46) |
|  | Weighted Score (0-10) | 0.92 (0.77, 1.11) |
| **Behavioral and Emotional Disorders** | 1 | 0.88 (0.67, 1.15) |
|  | 2 | 1.63 (1.19, 2.23)** |
|  | 3 | 1.46 (1.05, 2.03)* |
|  | ≥ 4 | 1.83 (1.47, 2.28)*** |
|  | Weighted Score (0-10) | 1.33 (1.23, 1.44)*** |
| **Personality Disorders** | 1 | 0.85 (0.37, 1.96) |
|  | 2 | 1.70 (0.77, 3.77) |
|  | 3 | 1.23 (0.50, 3.05) |
|  | ≥ 4 | 1.42 (0.76, 2.65) |
|  | Weighted Score (0-10) | 1.34 (1.10, 1.63)** |
| **Behavioral Syndromes Associated with Physiological Disturbances** | 1 | 0.89 (0.52, 1.52) |
|  | 2 | 1.33 (0.76, 2.35) |
|  | 3 | 0.82 (0.42, 1.63) |
|  | ≥ 4 | 1.46 (0.97, 2.19) |
|  | Weighted Score (0-10) | 1.18 (1.02, 1.35)* |

*Note.* ACEs = adverse childhood experiences. OR = odds ratio. CI = 95% confidence interval. 15 ACEs were analyzed for assessing counts and scores. Multivariate regressions controlled for sex and age. \* $p < .05$ , \*\* $p < .01$ , \*\*\* $p < .001$ .

Results in Table 5 are from a series of logistic regressions associating individual types of ACEs with neuropsychiatric outcomes. Across diagnoses, individual ACEs had substantial and significant associations with increased risk of psychiatric outcomes. For visualization and comparison, model coefficients were grouped into abuse and neglect (Figure 1B), care and family background (Figure 1C), and household history (Figure 1D) categories.

**Table 5.**
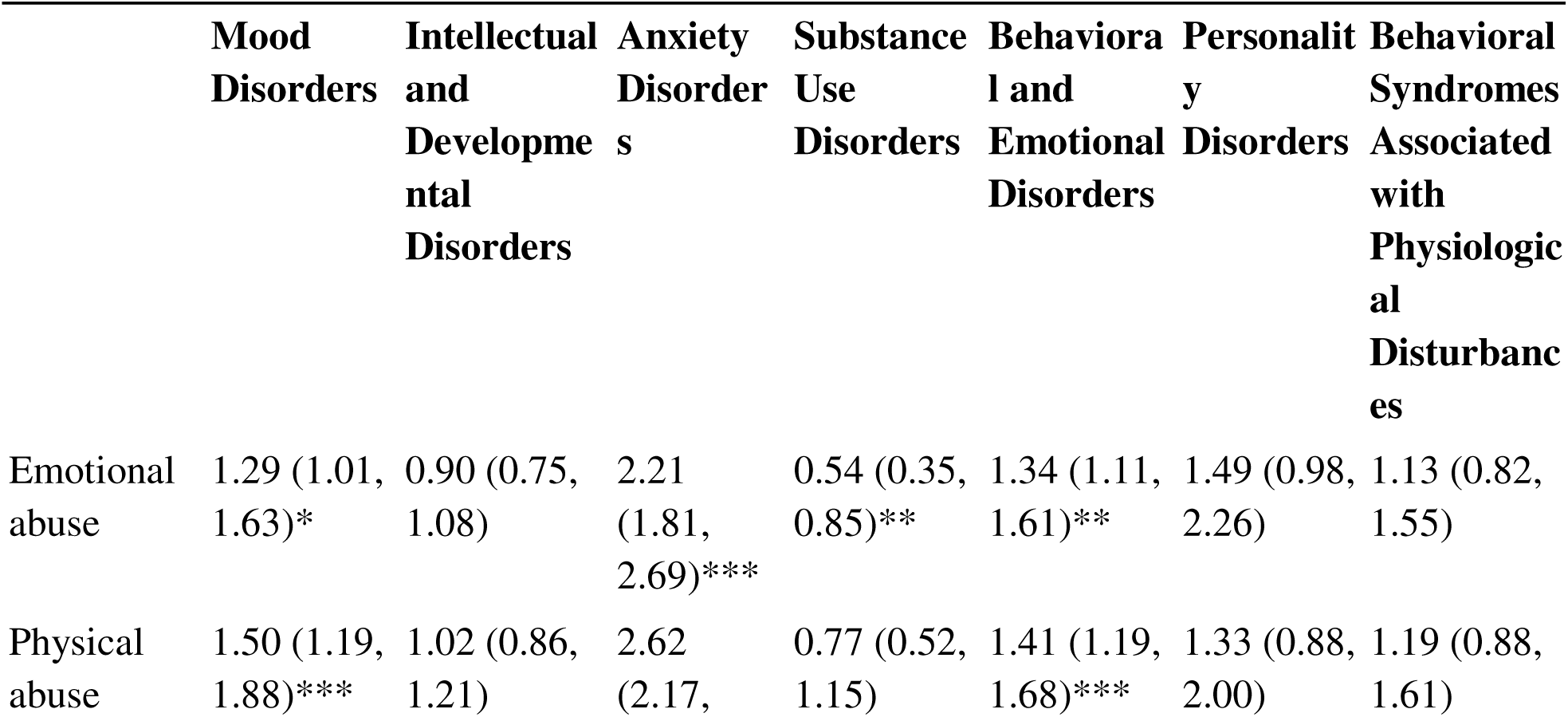
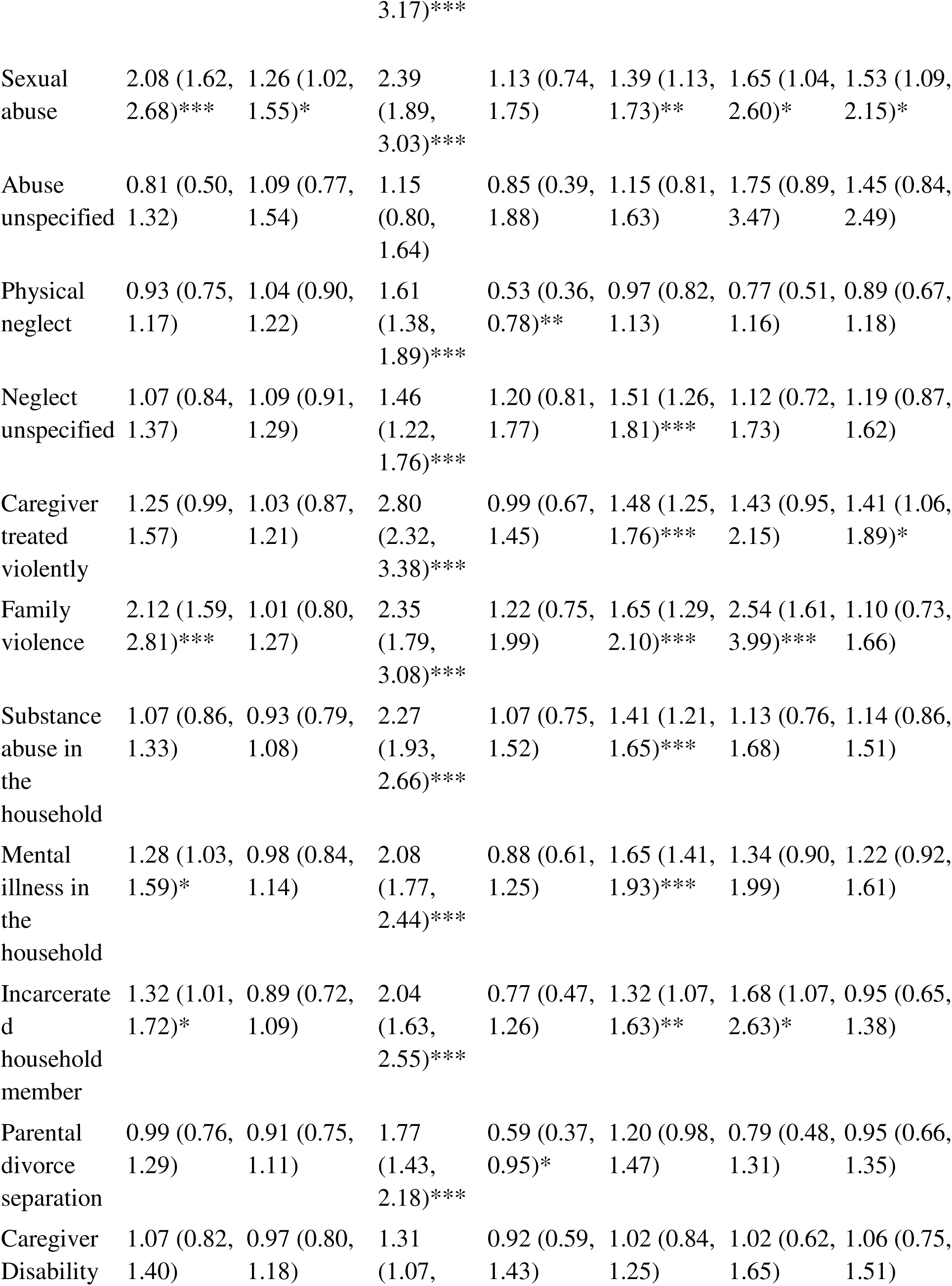
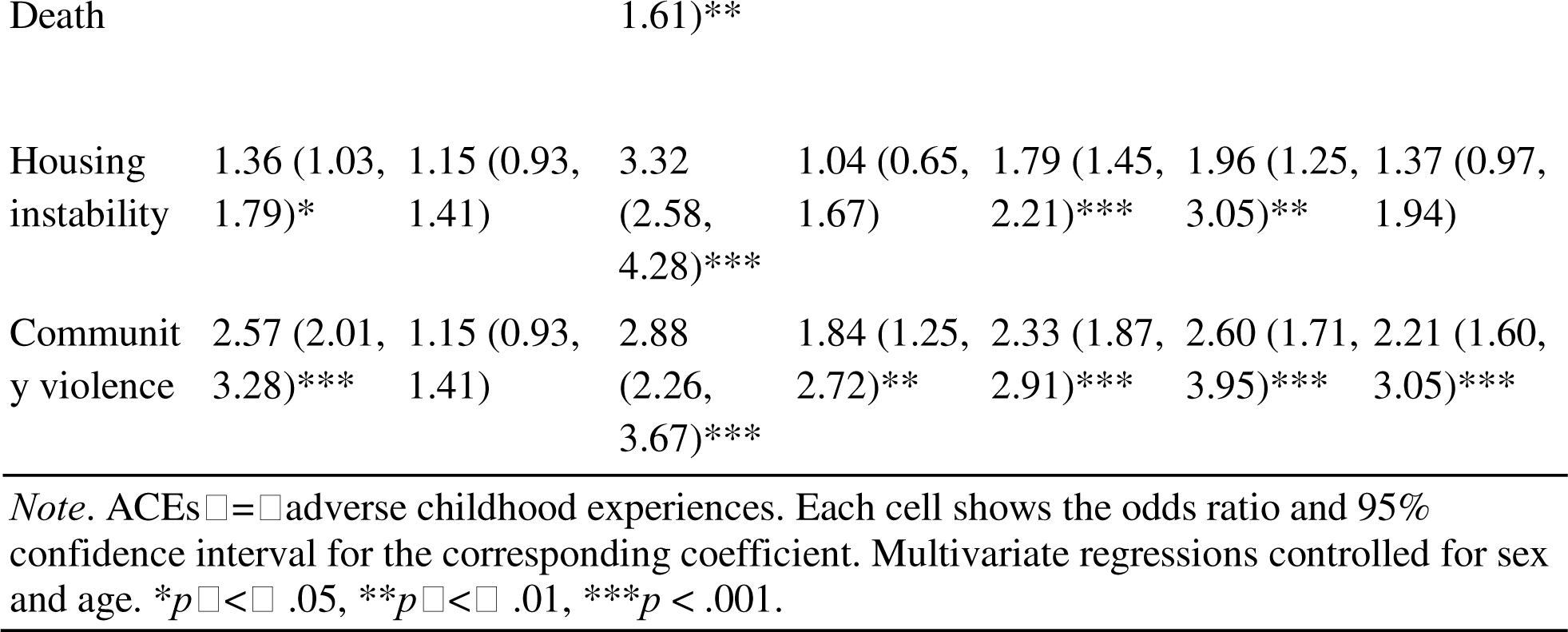
Associations between ACE types and neuropsychiatric outcomes.

In Figure 1, these plots highlight predictors across diagnostic outcomes and reveal diagnosis-specific patterns in effect direction and magnitude.

Results in Table 6 are from a series of logistic regressions associating individual elements of patient personal history data with neuropsychiatric outcomes. All personal history categories were strongly associated with increased risk of receiving psychiatric diagnoses. Personal history of self-injurious behavior (SIB) was significantly associated with every diagnostic category. Effect sizes and CIs for all personal history coefficients are visualized in Figure 2.

**Table 6.**
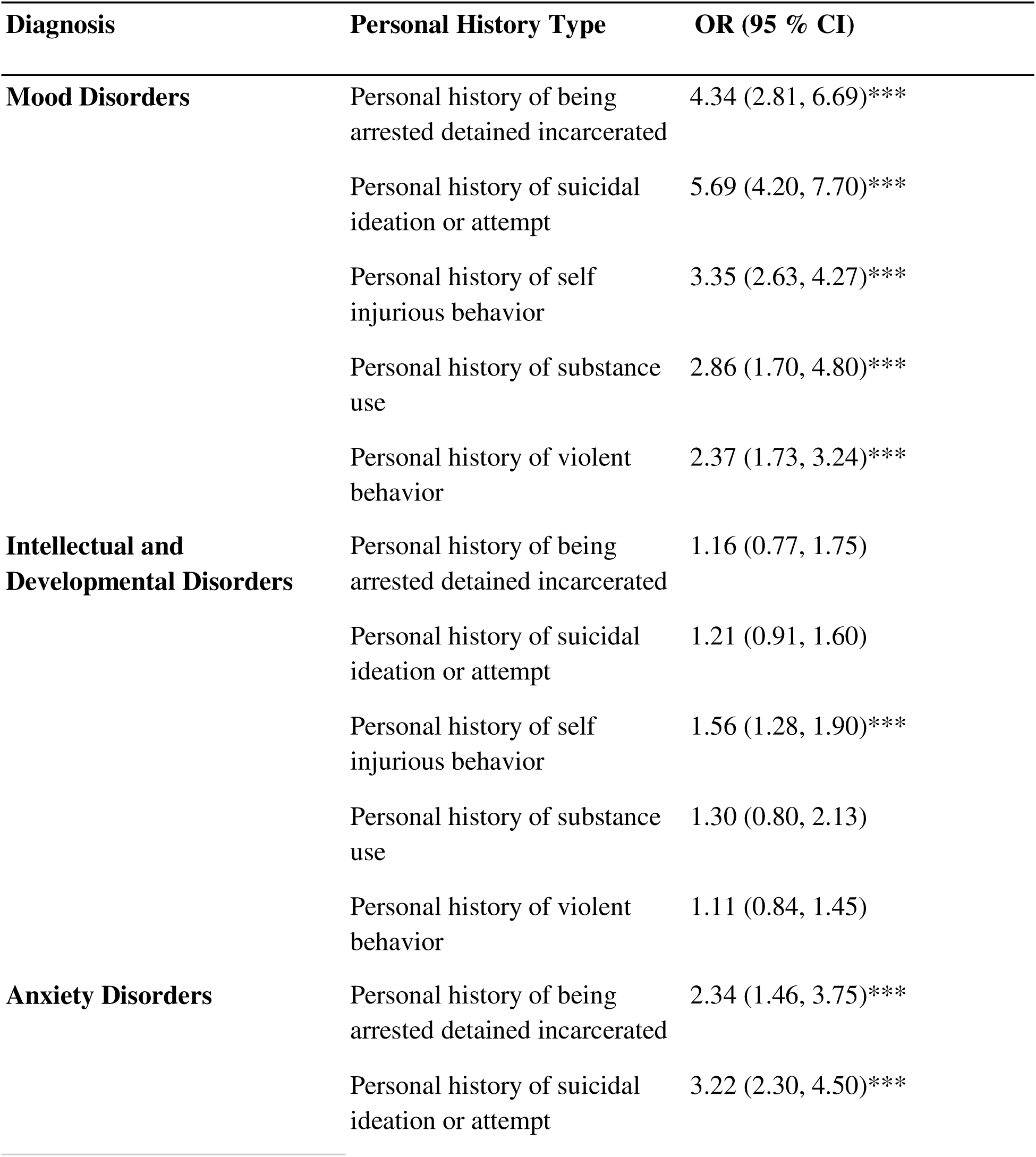
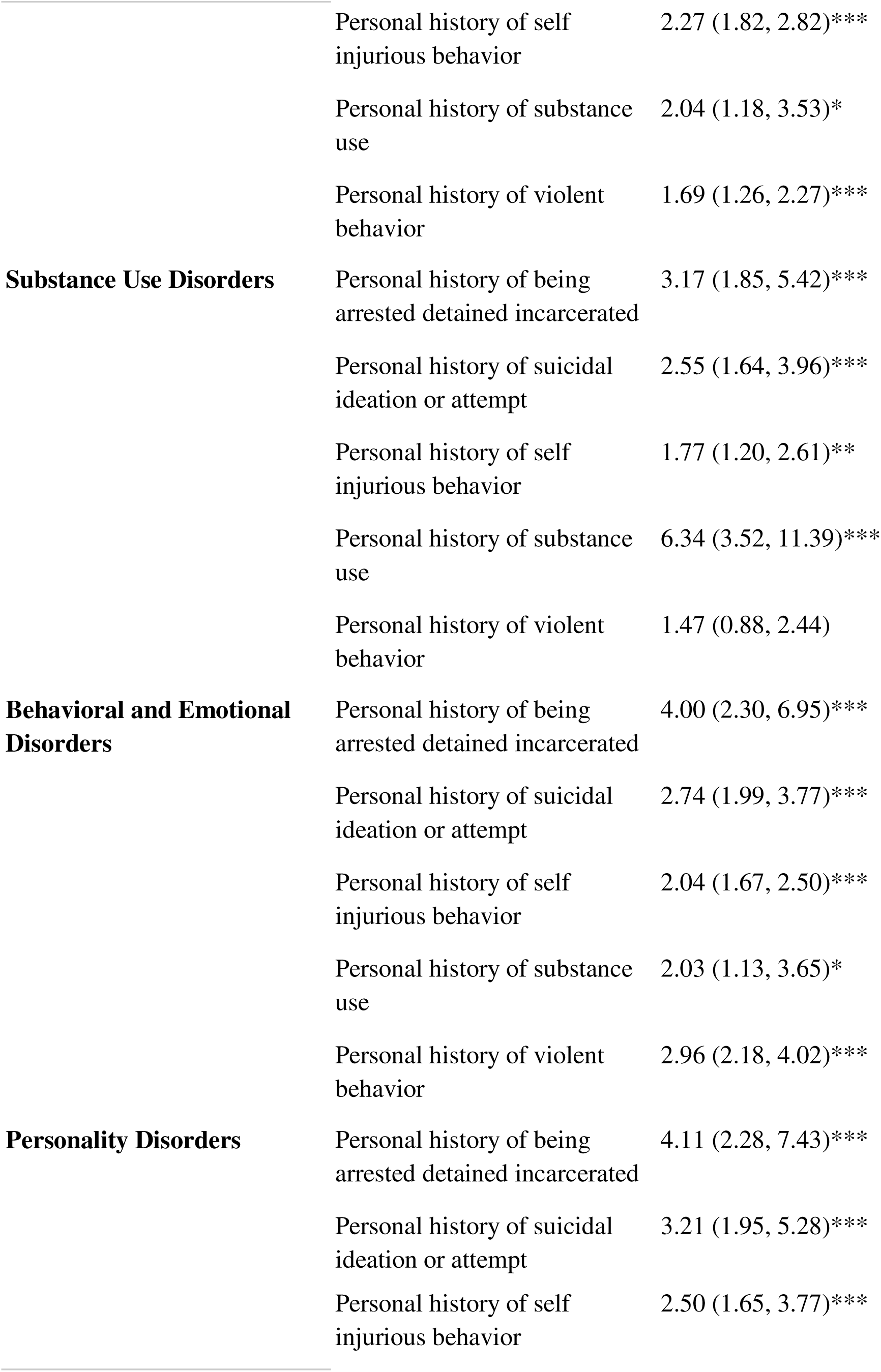
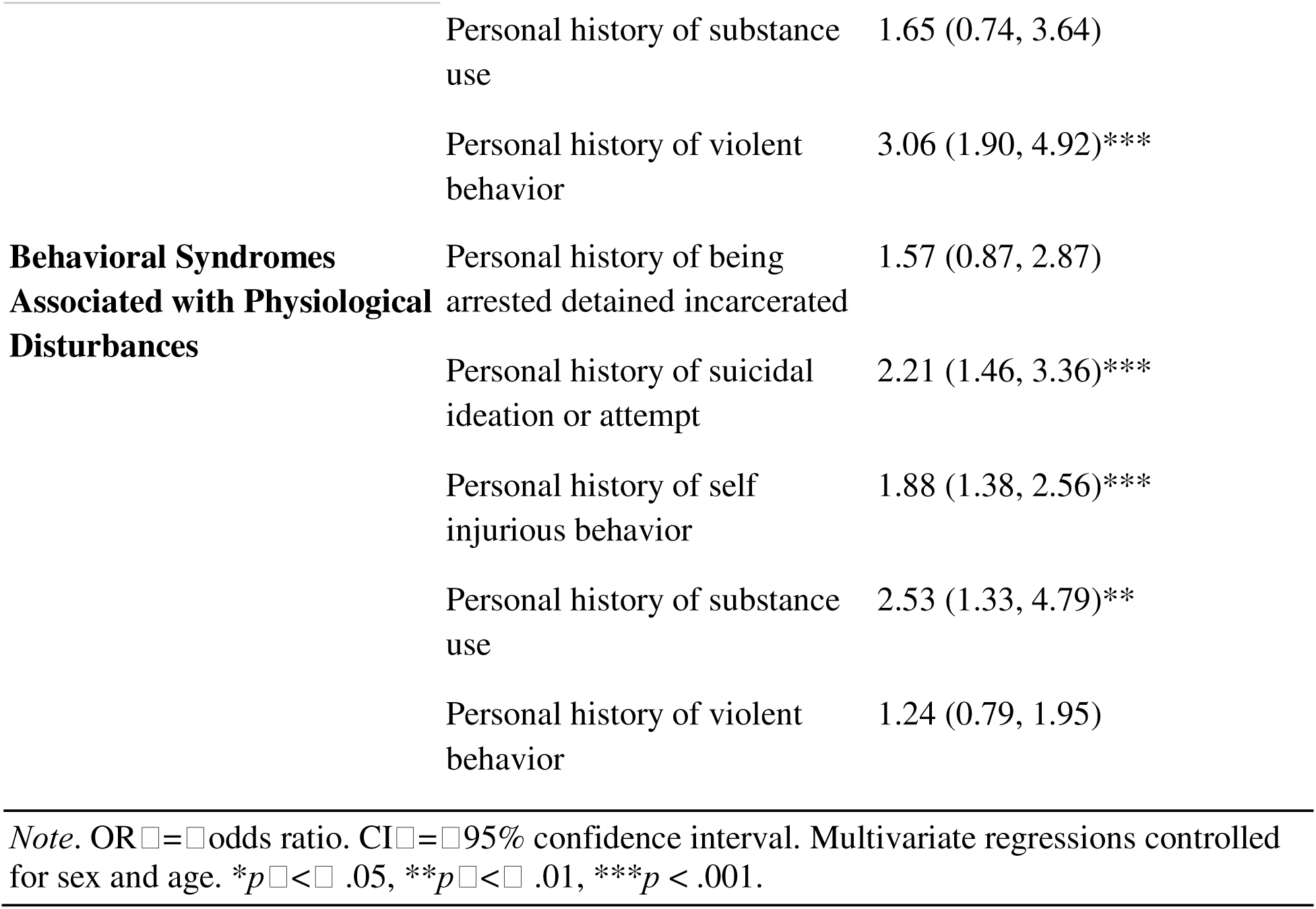
Associations between patient personal history and neuropsychiatric outcomes. Individual Risk Model.

| Diagnosis | Personal History Type | OR (95 % CI) |
| --- | --- | --- |
| **Mood Disorders** | Personal history of being arrested detained incarcerated | 4.34 (2.81, 6.69)*** |
|  | Personal history of suicidal ideation or attempt | 5.69 (4.20, 7.70)*** |
|  | Personal history of self injurious behavior | 3.35 (2.63, 4.27)*** |
|  | Personal history of substance use | 2.86 (1.70, 4.80)*** |
|  | Personal history of violent behavior | 2.37 (1.73, 3.24)*** |
| **Intellectual and Developmental Disorders** | Personal history of being arrested detained incarcerated | 1.16 (0.77, 1.75) |
|  | Personal history of suicidal ideation or attempt | 1.21 (0.91, 1.60) |
|  | Personal history of self injurious behavior | 1.56 (1.28, 1.90)*** |
|  | Personal history of substance use | 1.30 (0.80, 2.13) |
|  | Personal history of violent behavior | 1.11 (0.84, 1.45) |
| **Anxiety Disorders** | Personal history of being arrested detained incarcerated | 2.34 (1.46, 3.75)*** |
|  | Personal history of suicidal ideation or attempt | 3.22 (2.30, 4.50)*** |
|  | Personal history of self injurious behavior | 2.27 (1.82, 2.82)*** |
|  | Personal history of substance use | 2.04 (1.18, 3.53)* |
|  | Personal history of violent behavior | 1.69 (1.26, 2.27)*** |
| **Substance Use Disorders** | Personal history of being arrested detained incarcerated | 3.17 (1.85, 5.42)*** |
|  | Personal history of suicidal ideation or attempt | 2.55 (1.64, 3.96)*** |
|  | Personal history of self injurious behavior | 1.77 (1.20, 2.61)** |
|  | Personal history of substance use | 6.34 (3.52, 11.39)*** |
|  | Personal history of violent behavior | 1.47 (0.88, 2.44) |
| **Behavioral and Emotional Disorders** | Personal history of being arrested detained incarcerated | 4.00 (2.30, 6.95)*** |
|  | Personal history of suicidal ideation or attempt | 2.74 (1.99, 3.77)*** |
|  | Personal history of self injurious behavior | 2.04 (1.67, 2.50)*** |
|  | Personal history of substance use | 2.03 (1.13, 3.65)* |
|  | Personal history of violent behavior | 2.96 (2.18, 4.02)*** |
| **Personality Disorders** | Personal history of being arrested detained incarcerated | 4.11 (2.28, 7.43)*** |
|  | Personal history of suicidal ideation or attempt | 3.21 (1.95, 5.28)*** |
|  | Personal history of self injurious behavior | 2.50 (1.65, 3.77)*** |
|  | Personal history of substance use | 1.65 (0.74, 3.64) |
|  | Personal history of violent behavior | 3.06 (1.90, 4.92)*** |
| **Behavioral Syndromes Associated with Physiological Disturbances** | Personal history of being arrested detained incarcerated | 1.57 (0.87, 2.87) |
|  | Personal history of suicidal ideation or attempt | 2.21 (1.46, 3.36)*** |
|  | Personal history of self injurious behavior | 1.88 (1.38, 2.56)*** |
|  | Personal history of substance use | 2.53 (1.33, 4.79)** |
|  | Personal history of violent behavior | 1.24 (0.79, 1.95) |

*Note.* OR = odds ratio. CI = 95% confidence interval. Multivariate regressions controlled for sex and age. \* $p < .05$ , \*\* $p < .01$ , \*\*\* $p < .001$ .

**Figure 2:**
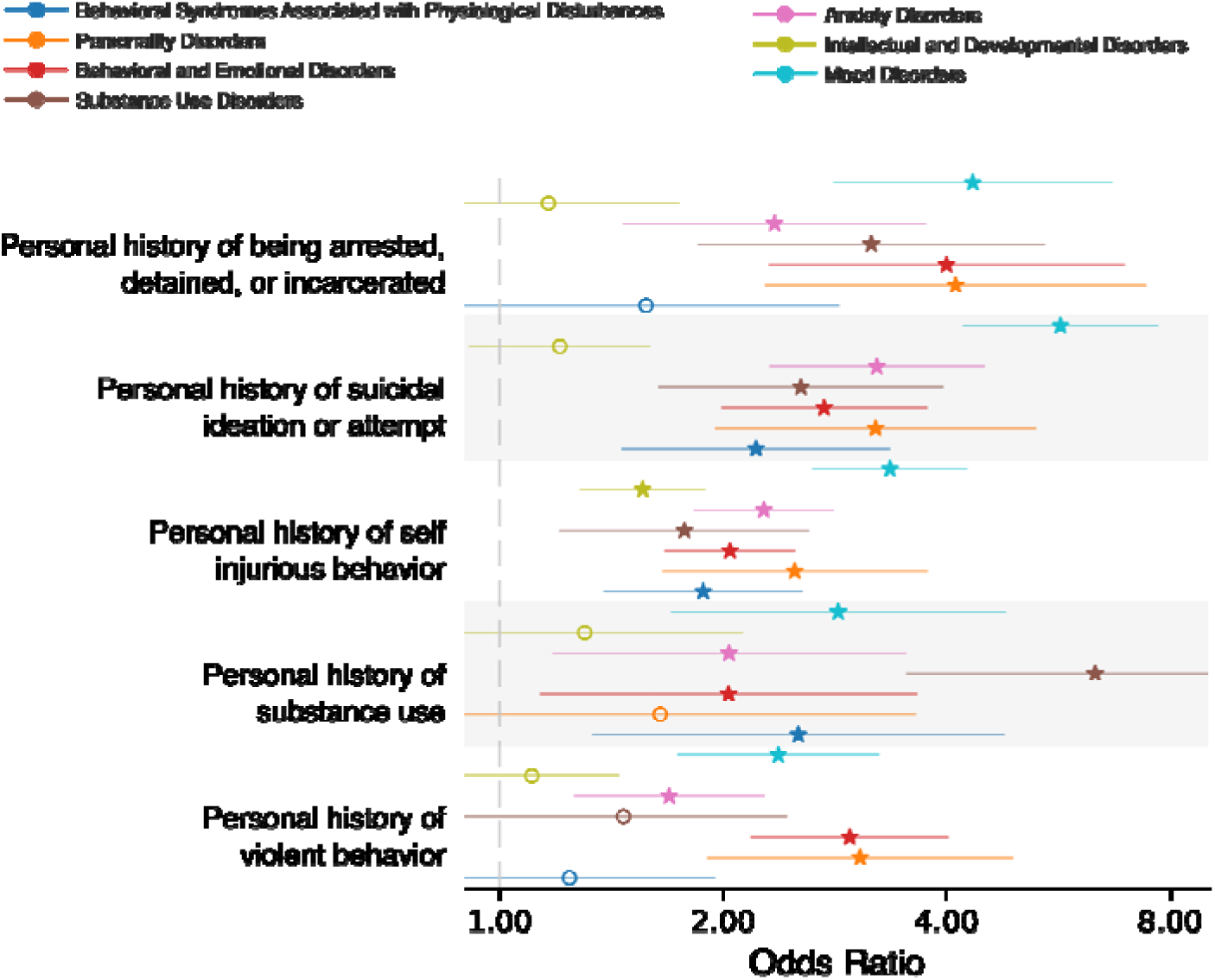
Forest plot of marginal effects of personal history categories on psychiatric and neurological indications. Each panel shows estimated effects and 95% confidence intervals for personal history categories across 7 diagnostic outcomes. Points indicate mean parameter estimates for odds ratios, and horizontal lines denote corresponding confidence intervals. Significant predictors are indicated by star markers. Outcomes for each personal history feature are grouped by row.

## Discussion

This study examined the relationship between formal psychiatric diagnoses and ACEs across multiple risk modeling approaches: threshold (dose-dependent), cumulative, and individual. This is the first known study to compare these frameworks to explore psychiatric outcomes among individuals recently adopted and/or currently in foster care who experienced ACEs. Our findings provide a significant contribution to current gaps in ACE research by utilizing a modeling approach for ACE count, score, and type, allowing researchers and clinicians to focus on ACE-specific risks between adversity and distinct psychiatric diagnoses.

### Threshold, dose-dependent

Previous studies establish a strong association between ACE count and increased odds of anxiety, depressive, and behavioral disorders.^5^ In our sample, ACE count showed a dose-dependent threshold pattern, with higher ACE counts associated with increased odds of Mood Disorders (F30–F39), Anxiety Disorders (F40–F48), and Behavioral and Emotional Disorders (F90–F98).

Mood disorders in childhood can disrupt development and are linked to elevated suicide risk.^7^ Consistent with prior work, our findings suggest that risk for Mood Disorders was most apparent at ≥4 ACEs. This aligns with previous evidence that higher ACE burden is associated with depression-related outcomes,^9^ and with findings that foster care–related adversities can be associated with depression risk.^14^ Given the substantial public health burden of depression, clarifying how adversity relates to clinically diagnosed mood disorders in high-risk youth remains important.

Anxiety disorders are common, often begin early, and may become chronic or relapsing.^7^ In our sample, increasing ACE counts were associated with higher odds of Anxiety Disorders, consistent with evidence that high ACE exposure is linked to substantially elevated anxiety risk in childhood.^15^ Finally, our threshold findings for Behavioral and Emotional Disorders are consistent with elevated rates of internalizing and externalizing disorders among youth in child welfare settings.^16^ Overall, greater childhood adversity was associated with higher risk across these psychiatric diagnostic domains.

### Cumulative ACE score

Cumulative weighted ACE scores have been consistently associated with poorer psychiatric outcomes.^17^ In our sample, higher cumulative ACE score was associated with increased odds across multiple diagnostic domains, including Mood Disorders, Anxiety Disorders, Behavioral and Emotional Disorders, Personality Disorders, and Behavioral Syndromes Associated with Physiological Disturbances. These findings align with prior evidence linking higher ACE burden to mood disorders and depression/bipolar outcomes,^17^ and to anxiety symptoms and diagnosis.^7^ Behavioral and Emotional Disorders also showed a graded association with cumulative score, consistent with elevated behavioral and emotional challenges among foster care youth with higher adversity exposure.^18^ Emerging longitudinal evidence further supports links between ACE exposure and later personality pathology, which is consistent with our observed association with the Personality Disorder domain. Finally, higher cumulative weighted ACE score was associated with Behavioral Syndromes Associated with Physiological Disturbances (e.g., sleep/eating disorders), consistent with evidence that adversity in foster/adopted populations is linked to sleep disturbances and related behavioral sequelae.^19^

### Individual

Individual risk modeling proposes that specific ACEs confer risk for the development of poor psychiatric outcomes. Our findings support individual risks for specific ACEs across several diagnosis profiles, where forms of violence and abuse had substantial impacts across almost all neuropsychiatric outcomes. Diagnosis domains of Anxiety Disorders, Behavioral and Emotional Disorders, and Mood Disorders were most commonly significant across ACE type, where all but one ACE type illustrated significant increased odds for the Anxiety Disorders diagnosis domain.

#### Emotional abuse

Emotional abuse was associated with increased odds of Mood, Anxiety, and Behavioral/Emotional Disorders, consistent with prior findings linking emotional abuse to mood disorder diagnoses.^4^ Emotional abuse was also associated with decreased odds of Substance Use Disorders.

#### Physical abuse

Physical abuse was associated with increased odds of Mood, Anxiety, and Behavioral/Emotional Disorders, aligning with evidence linking physical abuse to depressive, anxiety, and behavioral/conduct outcomes.^4^

#### Sexual abuse

Sexual abuse was associated with increased odds across most diagnostic domains, consistent with reports of strong associations with PTSD/anxiety and internalizing symptoms.^12^

#### Physical neglect

Physical neglect was associated with increased odds of Anxiety Disorders and decreased odds of Substance Use Disorders; prior work has linked neglect to anxiety, depression, and PTSD symptoms.^20^ The inverse association with Substance Use Disorders may reflect differences between clinician-diagnosed substance use disorders versus broader substance-use behaviors.

#### Neglect unspecified, (e.g. emotional, psychological, medical neglect)

Neglect (unspecified) was associated with increased odds of Anxiety Disorders and Behavioral/Emotional Disorders, consistent with evidence linking neglect to emotional and behavioral challenges.^21^

#### Caregiver treated violently

Witnessing a caregiver treated violently was associated with increased odds of Anxiety, Behavioral/Emotional Disorders, and Behavioral Syndromes Associated with Physiological Disturbances, consistent with prior work in foster care populations,^22^ and anxiety-related findings in maternal battery.^4^

#### Family violence

Family violence was associated with increased odds of Mood, Anxiety, Behavioral/Emotional, and Personality Disorders; prior work suggests family violence may disrupt attachment and socioemotional development, which may contribute to later personality pathology.^23^

#### Substance abuse in household

Household substance use was associated with three times the likelihood for Anxiety Disorders and Behavioral/Emotional Disorders, consistent with prior evidence linking household substance problems to internalizing and behavioral difficulties.^24^

#### Mental illness in the household

Household mental illness was associated with increased odds of Mood, Anxiety, and Behavioral/Emotional Disorders, consistent with prior studies.^3^

#### Incarcerated household member

Incarceration exposure was associated with increased odds of Mood, Anxiety, Behavioral/Emotional, and Personality Disorders, aligning with evidence linking household incarceration to later depressive/anxiety/stress-related diagnoses.^25^

#### Housing Instability

Housing instability was associated with increased odds across similar diagnostic domains including internalizing behaviors, depression, anxiety, PTSD, and increased risk of externalizing behaviors that can escalate to later behavioral, emotional, and personality disorders.^26^

#### Parental divorce separation

Parental divorce or separation held significance for increased odds of Anxiety Disorders and decreased odds in Substance Use Disorders. Similar to findings in Bohman et al (2017) where individuals that experienced parental divorce or separation had a higher likelihood of anxiety disorders into adulthood compared to non exposed individuals.^27^

#### Caregiver Disability Death

Caregiver disability/death was associated with increased odds of Anxiety Disorders, consistent with evidence linking caregiver loss/disruption to internalizing outcomes.^22^

#### Community violence

Studies show exposure to violent adverse events can dramatically impact an individual’s internalizing and externalizing behavior regulation.^28^ In contrast, our findings show that community violence was significant for increased odds in all diagnosis domains besides Intellectual and Developmental Disorders. Community violence is not commonly explored in current ACE research, but similar to family violence, it can impact how an individual navigates personal and community relationships, which supports our findings for this ACE across almost all measured diagnosis domains.

Overall, ACE type adds clinically useful specificity beyond ACE count, highlighting especially strong vulnerability associated with abuse and violence exposures.

##### Personal History

Personal history categories were associated with increased risk of receiving a diagnosis across psychiatric domains. Across all personal history categories, we saw strong significance for domains of Mood Disorders, Anxiety Disorders, and Behavioral and Emotional Disorders.

#### Personal history of arrested, detained, or incarcerated

Personal history of incarceration, detainment, or arrest refers to individuals placed in detention or involuntary confinement in police, emergency, or psychiatric inpatient settings. Prior research shows that foster care placement is associated with later adult criminal justice involvement and adverse psychiatric outcomes.^29^ Our findings align with this literature, demonstrating significant associations with Mood, Anxiety, Substance Use, Behavioral and Emotional, and Personality Disorders. Youth involved in the justice system are disproportionately affected by health conditions: 60-70% of detained or incarcerated adolescents meet criteria for at least one psychiatric disorder, compared to approximately 20% in the general population.^30^ Moreover, 50-75% of detained youth later enter the adult criminal justice system, and studies found that in 2014 only 43% of juvenile detention centers provided therapeutic services, potentially compounding long-term risk.^30^ Together, these findings highlight significant mental health risks associated with early justice involvement.

#### Personal history of being suicidal ideation or attempt

Personal history of suicide attempt or suicidal ideation was significantly associated with Mood Disorders, Anxiety Disorders, Substance Use Disorders, Behavioral and Emotional Disorders, Personality Disorders, and Behavioral Syndromes Associated with Physiological Disturbances, coinciding with previous research linking these disorders to suicide.^31^

#### Personal history of self-injurious behavior

SIB was significantly associated with increased odds across all diagnostic domains in this study. Interestingly, SIB was the only cluster in our study to be significantly linked to Intellectual and Developmental Disorders, which aligns with previous findings that those with intellectual disabilities were almost five times more likely to engage in SIB than the general population.^32^

#### Personal history of substance use

Personal history of substance use is confirmed by individuals through self-report or a toxicology screening before their first AMC appointment. This category was found to be significant with Mood Disorders, Anxiety Disorders, Substance Use Disorders, Behavioral and Emotional Disorders, and Behavioral Syndromes Associated with Physiological Disturbances, which is supported by previous research findings linking substance use and its severity with mental health disorders.^33^ This, as well as findings linking youth substance use with a mental health diagnosis, provides evidence that the presence of mental health disorders can exacerbate substance use.^34^

#### Personal history of violent behavior

This category was found to be significantly associated with Mood Disorders, Anxiety Disorders, Behavioral and Emotional Disorders, and Personality Disorders, which is supported by previous findings.^35^ For individuals in foster care, bullying also appears to be much more common, with a prevalence ranging from 55-73% depending on age, race/ethnicity, and gender.^36^ These acts of violence and bullying can have cascading negative effects in the communities or family units in which they occur.

While disorders like anxiety, depression, Attention-Deficit/Hyperactivity Disorder (ADHD), Post-Traumatic Stress Disorder (PTSD), and oppositional defiant disorder are implicated in all personal history findings, it is uncertain if they are predictors of the behavior described or if the affected individual may have difficulty engaging with programs that reduce these behaviors, showcasing the necessity for timely intervention.

## Conclusion

ACEs during critical adolescent neurodevelopment may be associated with heightened vulnerability among youth with foster care/adoption histories, contributing to impairments in social, emotional, and behavioral functioning that often present as internalizing (depression, anxiety) and externalizing symptoms (disruptive behavior, aggression).^13^ Timely identification of elevated ACE exposure is essential to enable early, trauma-focused interventions and tailored supports for social, emotional, and behavioral challenges in this population. Considering high-risk subgroups (e.g., adolescents in foster care) may further inform targeted prevention strategies.

Our study strengthens the evidence base by assessing ACE count, score, and type in relation to psychiatric diagnoses. Comparing cumulative, threshold, and individual risk models provides ACE-specific estimates of psychiatric risk and clarifies how adversities cluster across diagnostic categories. Additional strengths include a large UMN-AMC sample with standardized ACE data and psychiatric diagnoses, with adjustment for sex and age at visit. Future work should examine synergistic effects among commonly co-occurring ACEs.

In conclusion, ACE exposure was associated with increased odds across multiple psychiatric diagnostic domains. These findings support both categorical and continuous ACE modeling and underscore the importance of early, trauma-informed prevention and intervention for foster care and adopted individuals.

## Limitations

ACE data from the UMN-AMC come from first-person (e.g., biological parent report) and secondary (foster families, kinship caregivers, social workers) sources that confirm or describe exposure. Because secondary sources are limited, ACE exposure may be underreported. ACEs also vary in frequency, duration, intensity, and individual impact. For community and family violence variables, patient-perpetrated violence was prioritized; some youth who perpetrated violence may also have experienced violence. Future research should incorporate greater ACE dimensionality to clarify associations with psychiatric disorders. Many families seek UMN-AMC evaluations to access educational and disability resources; thus, youth with existing behavioral or health concerns may be more likely to present, potentially inflating psychiatric outcome rates. The Substance Use Disorder (SUD) diagnostic domain saw inverse odds for the ACE types of emotional abuse, physical abuse, and parental separation, which could be explained by variation in substance use and formal SUD diagnosis, age and timing of SUD development, and possible protective barriers like increased engagement in services and monitoring that could reduce progression to diagnosable SUDs. Future studies with age-matched controls are needed to improve generalizability.

## Future Directions

As our study focused on adversity and negative mental health outcomes, examining how protective factors of individuals with ACE history alongside resilience pathways could build on our findings. Additional exploration of detailed ACE variables incorporating domains of SES background, exposure and or perpetration of violence may provide additional clarity between specific adversities and psychiatric disorders. Lastly, as our study population consisted of individuals commonly located in the MN metro, additional research corroborating these models using different study populations outside of MN would support our findings.

## Data Availability

All data produced are available online at https://conservancy.umn.edu/browse/author?value=Randolph,%20Anita&bbm.return=1

## Declarations

### Ethics approval and consent to participate

This study was conducted under the oversight of the UMN Institutional Review Board. All procedures were in compliance with federal regulations, institutional policies, and ethical guidelines for human research. No informed consent was required for this retrospective chart review.

### Consent for publication

Not applicable

### Availability of data and materials

De-identified participant data—including baseline demographics, prenatal exposures, psychiatric diagnoses, and more, are available for analysis in the <u>Data Repository for the University of Minnesota (DRUM)</u>. The two de-identified datasets used in this study Building Understanding of Transitions & Trauma in Early Formative Life Years (BUTTERFLY) and Clinical Outcomes & Medical Profiles of Children in Adoption & Statecare Settings (COMPASS) can be found at these respective links: https://hdl.handle.net/11299/279102; https://hdl.handle.net/11299/279103 This is protected by an attribution noncommercial sharealike 4.0 international license. You must give appropriate credit, provide a link to the license, and indicate if changes were made. You may not in any way that suggests the licensor endorses you or your use. You may not use the material for commercial purposes. If you remix, transform, or build upon the material, you must distribute your contributions under the same license as the original. You may not apply legal terms or technological measures that legally restrict others from doing anything the license permits.

### Authors’ contributions

AR and MF conceived the project. AR secured funding. AR, LC, and SA supervised data capture, entered data manually, and validated data of enrolled patients. LC and SA provided access to the dataset. ED-VR, AK, RB, and AR curated the data. EK, MF, and AR supervised initial data entry and analysis. ED-VR and AH supervised data analysis and led data clustering and analysis with outcomes, and data visualization. ED-VR and AH analysed data, including data modelling and statistical analysis. All authors had access to the raw data. ED-VR and LC wrote the initial draft of the manuscript. LC, SA, CS, SS, and AR wrote the final manuscript. All the authors reviewed the final version of the manuscript and had full access to all data reported in the study. ED-VR, AH, AK, and RB verified the data.

## Acknowledgements

We thank all the staff of the University of Minnesota, Adoption Medicine Clinic. We would also like to thank the individuals that helped lay the initial methodological framework for this data, as well as helped with data entry and quality control: Taylor Rawstern, Jada Loleng, Jonathan Lehman, Jade Roghair

## List of Abbreviations

ACEs: Adverse childhood experiences
UMN-AMC: University of Minnesota Adoption Medicine Clinic
EHRs: Electronic Health Records
CCWA: Comprehensive Child Wellness Assessments
BPIC: Best Practices Integrated Informatics Core
SIB: Self-Injurious Behavior
SUD: Substance Use Disorder

